# Rural Roads to Cognitive Resilience (RRR): A prospective cohort study protocol

**DOI:** 10.1101/2024.10.13.24315411

**Authors:** Lilah M. Besser, Lisa Wiese, Diane J. Cook, Janet Holt, Sheryl Magzamen, Bryan Minor, Diana Mitsova, Juyoung Park, Olivia Sablan, Madeleine Tourelle, Christine Williams

## Abstract

**Background:** Ambient air pollution, detrimental built and social environments, social isolation (SI), low socioeconomic status (SES), and rural (versus urban) residence have been associated with cognitive decline and risk of Alzheimer’s disease and related dementias (ADRD). Research is needed to investigate the influence of ambient air pollution and built and social environments on SI and cognitive decline among rural, disadvantaged, ethnic minority communities. To address this gap, this cohort study will recruit an ethnoracially diverse, rural Florida sample in geographic proximity to seasonal agricultural burning. We will (1) examine contributions of smoke-related fine particulate matter (PM_2.5_) exposures to SI and cognitive function; (2) determine effects of built and social environments on SI and cognitive function; and (3) contextualize SI and cognitive function among residents from different ethnoracial groups during burn and non-burn seasons.

**Methods:** We will recruit 1,087 community-dwelling, dementia-free, ≥45-year-olds from five communities in Florida’s Lake Okeechobee region. Over 36 months, participants will complete baseline visits to collect demographics, health history, and health measurements (e.g., blood pressure, body mass index) and 6-month follow-ups assessing cognitive function and social isolation at each visit. A subsample of 120 participants representative of each community will wear smartwatches to collect sensor data (e.g., heart rate) and daily routine and predefined activities (e.g., GPS-captured travel, frequent destinations) over two months. Ecological momentary assessments (EMA) (e.g., whether smoke has bothered participant in last 30 minutes) will occur over two months during agricultural burning and non-burning months. PurpleAir monitors (36 total) will be installed in each community to continuously monitor outdoor PM_2.5_ levels.

**Ethics and expected impact:** This study received Florida Atlantic University’s Institutional Review Board approval and will require participant informed consent. We expect to identify individual- and community-level factors that increase the risk for SI and cognitive decline in a vulnerable rural population.

## INTRODUCTION

Alzheimer’s disease and related dementias (ADRD) comprise multiple neurodegenerative diseases with differing neuropathological origins, including Alzheimer’s disease, Lewy body disease, and frontotemporal degeneration [1–3]. Over time, the neuropathological burden of these diseases leads to dementia, which is the “loss of memory, language, problem-solving and other thinking abilities that are severe enough to interfere with daily life” [4]. ADRD risk increases with age and is 1.5 to 2 times higher among Black and Hispanic individuals, who are more likely to have lower educational attainment and chronic conditions (e.g., diabetes, hypertension, cardiovascular disease) that are known risk factors for ADRD [5, 6]. Social determinants of health (SDOH), which are the conditions where people live, work, worship, play, and age, have been increasingly implicated in increasing ADRD risk, particularly for minoritized and socioeconomically disadvantaged communities [7–9]. For instance, Black and Hispanic older adults tend to live in neighborhoods with greater area deprivation and lower greenspace access, and these factors have been associated with greater risk of cognitive decline and dementia in numerous studies [10–14].

In addition, both air pollution and social isolation are key SDOH that have been named by the Lancet Commission as ADRD risk factors [6, 15], and exposures to ambient air pollution and social isolation are more prevalent in historically disadvantaged communities [16, 17]. Ambient air pollution, including particulate matter of <2.5μm in size (PM_2.5_), has been shown to increase morbidity and mortality and ADRD risk, particularly among urban-dwelling Hispanic and Black individuals [18–23]. The preponderance of air quality health effects studies in the U.S. have been conducted using U.S. Environmental Protection (EPA) Agency Air Quality System data and have focused largely in urban areas where monitors are located. Agricultural burns, like wildfire smoke, have smoke plumes with large spatiotemporal gradients which frequently impact sparsely populated areas with limited extant monitoring. Yet, agricultural fires, unlike wildfires, have constrained size, intensity, and smoke production, thereby testing the limits of detection by satellite. A major advantage of studying agricultural burns is that in contrast to wildfires, agricultural fires follow a predictable schedule in circumscribed spatial areas, enabling ground monitoring. However, studies on the health effects from rural agricultural fires which disproportionately impact Black and Hispanic communities are limited [23].

Social isolation, recognized as a significant threat to public health by the U.S. Surgeon General, is characterized by the lack of social engagement, supportive social networks, and/or participation in social activities [24]. Numerous studies have linked social isolation to poorer cognitive function and ADRD risk [25–31]. For example, 70-year olds from the Lothian Birth Cohort who lived alone had slower processing speed and those who reported greater loneliness had worse overall cognitive function [27].

While sufficient evidence is available to implicate social isolation in increasing ADRD risk, the published studies are primarily limited to urban and White populations, with little known about how these factors impact rural and ethnoracially diverse populations that are most vulnerable to developing ADRD.

Neighborhood social and built environments are SDOH that have also been associated with cognitive function and ADRD risk [8, 10, 11, 32, 33]. For instance, neighborhood social environments, such as living in areas with greater area deprivation, greater psychosocial disorder (e.g., crime), and ethnoracial segregation, have been associated with worse ADRD-related outcomes in prior studies [13, 34, 35]. Built environments are manmade physical environments including but not limited to transportation and pedestrian networks, social and walking destinations (shops, libraries, banks, restaurants), parks and other greenspaces, and health care facility access. A systematic review of studies on various built environment factors found the strongest evidence to date was for associations between greater walking/social destinations and more greenspace in the neighborhood and better cognitive function and lower dementia risk [32]. However, similar to the studies focused on either social isolation or air pollution exposure and ADRD risk, few studies of neighborhood environments and ADRD have centered on rural, ethnoracially diverse older adults.

Rural residence (versus urban) has been associated with higher incidence or prevalence of ADRD [36, 37]. Some reasons for this may be lower educational attainment and lower access to health care and other resources that help maintain brain health into later life [38–40] However, the relationship between rural/urban residence and ADRD risk is not straightforward given the differences in access to care and diagnostic services in rural areas that may lead to underdiagnosis of ADRD, in addition to the significant differences in the ethnoracial groups living in rural areas depending on U.S. region [41]. Overall, few studies have been conducted on risk factors for ADRD among ethnoracially diverse rural populations in the U.S.

Altogether, the extant literature suggests that several SDOH at the environmental (i.e., air pollution, neighborhood social and built environments, rural residence) and individual level (i.e., ethnoracial group and social isolation) increase ADRD risk. However, no known studies have investigated how all these factors considered together contribute to cognitive decline in older adults, particularly those living in rural areas with varying seasonal levels of PM_2.5_ due to agricultural burning. To address this significant scientific gap, the Rural Roads to cognitive Resilience (RRR) cohort study will recruit a sample of ethnoracially diverse (predominantly Black and Hispanic), socioeconomically disadvantaged older adults living in a rural agricultural area surrounding Lake Okeechobee in South Central Florida. The study, which targets a region beset by a combination of the aforementioned and other unique ADRD risks, aims to (1) examine the contribution of agricultural smoke-related PM_2.5_ exposures to social isolation and cognitive function, (2) determine the effects of the built (e.g., park space) and social environment (e.g., neighborhood disadvantage) on social isolation and cognitive function, and (3) contextualize social isolation and cognitive function among residents from different ethnoracial groups using ecological momentary assessment (EMA) and smartwatch sensor-derived behavior models with a subsample of 120 stratified by Lake O communities during agricultural burn and non-burn seasons.

## MATERIALS AND METHODS

### Setting and design

#### Study summary

Rural Roads to cognitive Resilience (RRR) is a longitudinal, observational cohort study that will broadly collect data on ambient air pollution exposure, neighborhood built and social environments, social isolation, and cognitive functioning among participants 45 and older living in rural communities surrounding Lake Okeechobee, Florida. The study is comprised of two data collection efforts: (i) a 36-month study of ∼1,000 individuals to investigate social isolation and cognitive function may be affected by ambient PM_2.5_ and neighborhood built/social environments, and (ii) a smartwatch study subsample of ∼120 participants recruited from the 36-month study.

#### Setting

The Everglades Agricultural Area in South Central Florida is the largest sugarcane-producing region in the U.S. from ∼October to May each year, sugar companies employ farmworkers to prepare 400,000 acres of fields for harvesting by burning the sugarcane outer leaves. The population surrounding Lake Okeechobee (Lake O) demonstrates a multitude of disparities that are risk factors for ADRD. For example, in the town of Belle Glade, 40% of the population did not graduate from high school, health literacy is equivalent to 7th grade, 91% are minoritized ethnic groups, the area has limited healthcare access, and poverty affects 41% of the population [42]. In addition, during burns, residents surrounding Lake Okeechobee describe experiences of stench, eye irritation, feeling tight in the chest, and outdoor areas covered in soot, preventing outdoor social engagement [43]. Typically, the background PM_2.5_ levels in the Lake O region are relatively low, but increases of up to 2μg/m^3^ occur during agricultural burning season [44]. Altogether, Lake O provides a fitting setting to study the impact of air pollution on diverse, rural, and high-risk communities.

##### Specific aims

Aim 1. Examine the contribution of particulate matter <2.5μm (PM_2.5_) exposures to social isolation and cognitive function (CF), through multilevel growth modeling.

- *Aim 1a.* Examine baseline levels and growth trajectories of SI and CF associated with changes in PM_2.5_ during agricultural burn and non-burn seasons over a 36-month period, accounting for individual- and neighborhood-level characteristics.
- *Aim 1b.* Examine the effect modification of race/ethnicity on the relationship of PM_2.5_ on baseline levels and growth trajectories of SI and CF, accounting for individual- and neighborhood-level characteristics.

Aim 2. Determine the effects of the built environment (e.g., retail destinations, park space) and social environment (e.g., crime, SES) on SI and CF through linear mixed modeling.

- *Aim 2a.* Examine SI and CF associated with the built and social environment over a 36-month period (baseline, and FY 1,2,3), controlling for individual- and other neighborhood-level characteristics.
- *Aim 2b.* Explore the effect modification of race/ethnicity on the relationship of the built and social environment with SI and CF, in view of their educational and social disadvantage.

Aim 3. Contextualize SI and CF among residents from different ethnoracial groups using EMA and sensor-derived behavior models during burn and non-burn seasons in a subsample of 120 ethnoracially diverse participants, stratified by Lake O communities.

- *Aim 3a.* Examine residents’ momentary changes in behavioral markers, mood, and cognitive performance, associated with changes in air quality and the social and built environment.
- *Aim 3b.* Characterize differences among ethnoracial groups and between women and men in their momentary changes in behavioral markers, mood, and CF performance in response to changes in air quality and the social and built environment.

#### Enrollment

RRR will recruit a sample of approximately 1,000 individuals located in Hendry, Glades, Okeechobee, and Palm Beach counties. More specifically, participants will be enrolled from 4 rural communities (i.e., Clewiston, Pahokee/Canal Point, Okeechobee, Belle Glade/South Bay) and 1 suburban/urban community (i.e., Royal Palm Beach). The communities were chosen to provide diversity in ethnoracial and socioeconomic groups and socioeconomic status (higher Area Deprivation Index score: more deprived neighborhood) and sufficient variation in social and built environments at the census block group level to successfully complete the study aims (Figures 1-4).

**Figure 1.**
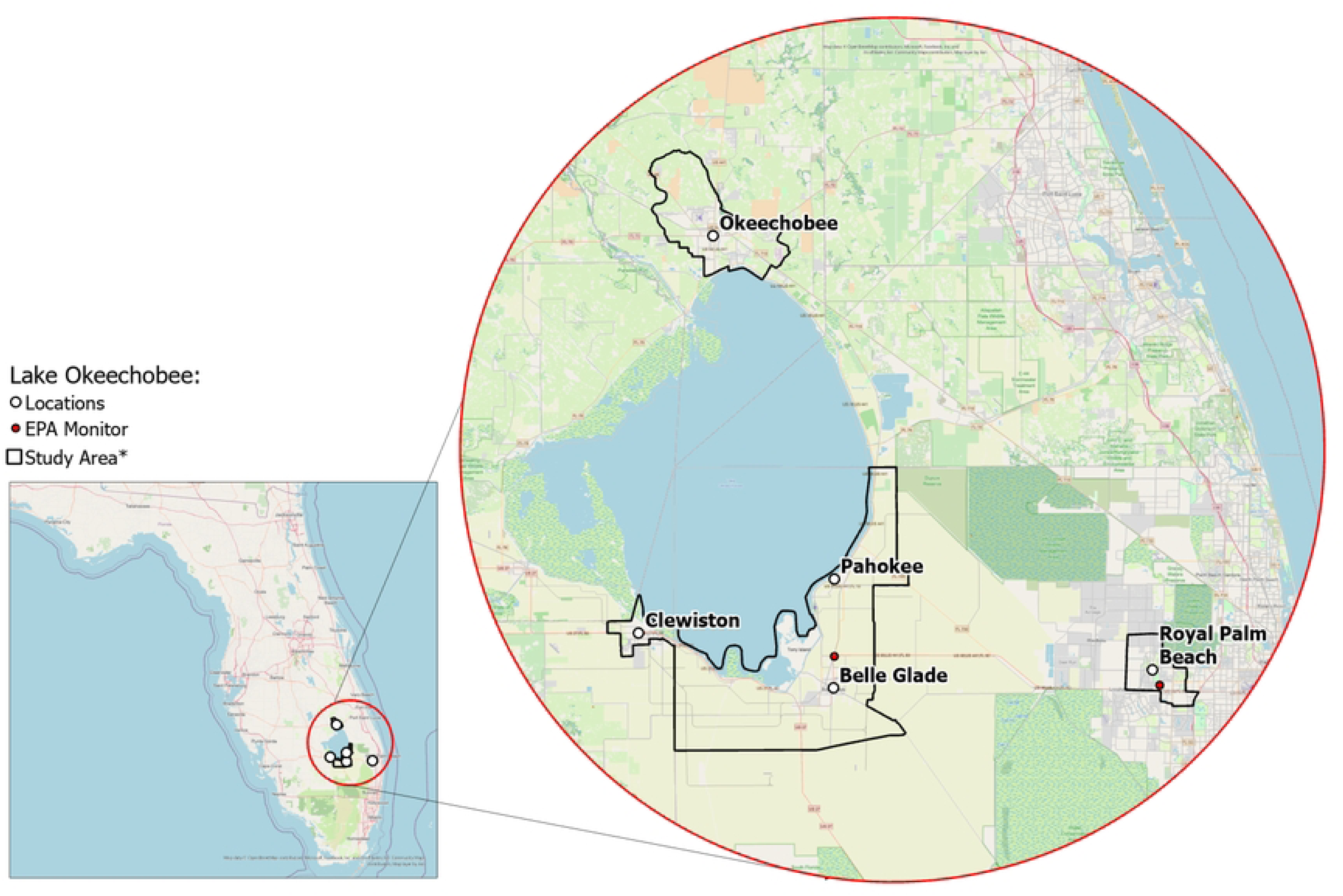
Florida communities included in the study. * Exact locations/census block groups included will depend on recruitment.

**Figure 2.**
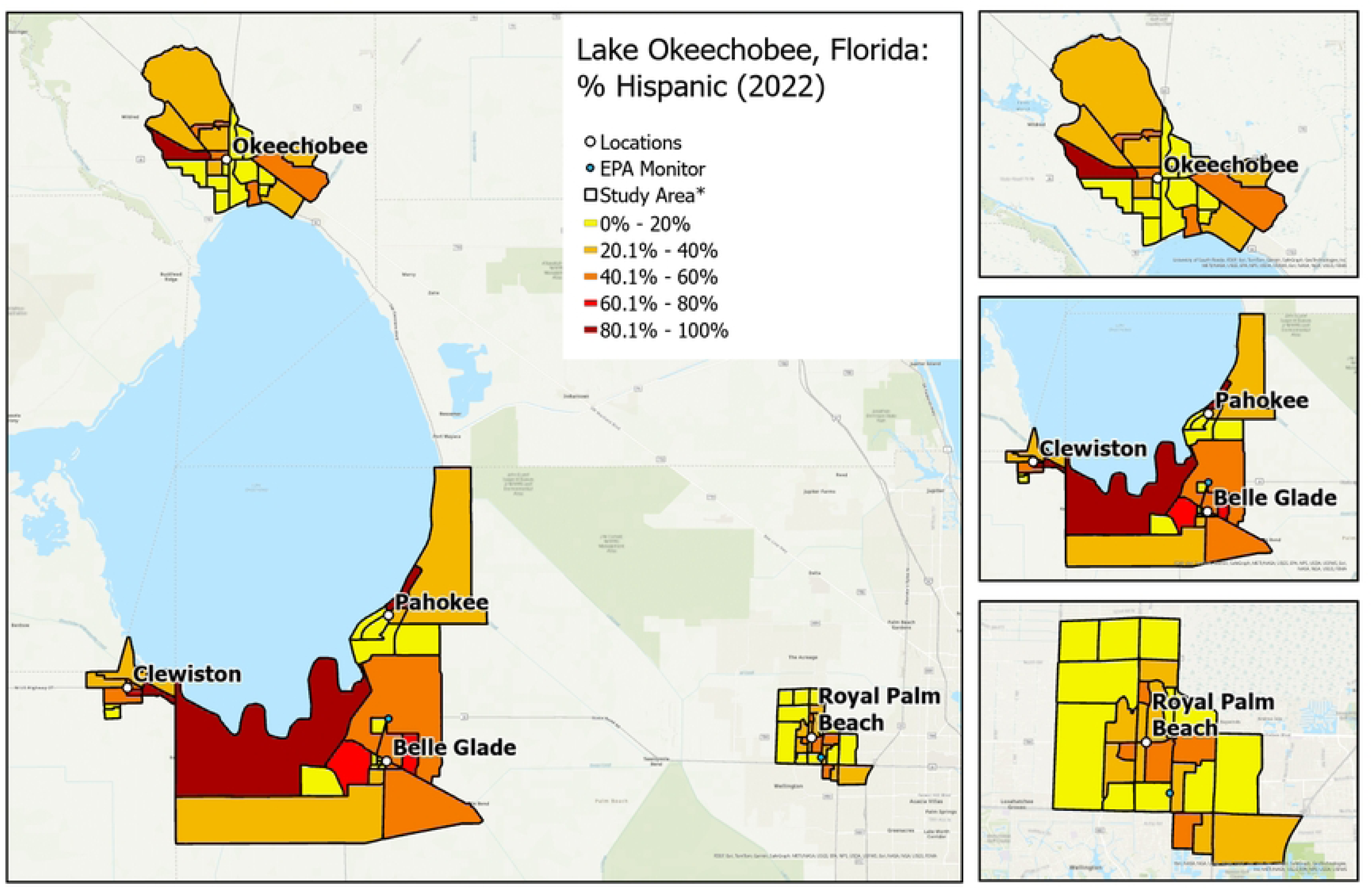
Variation in neighborhood percentage of Hispanic residents by community. * Exact locations/census block groups included will depend on recruitment.

**Figure 3.**
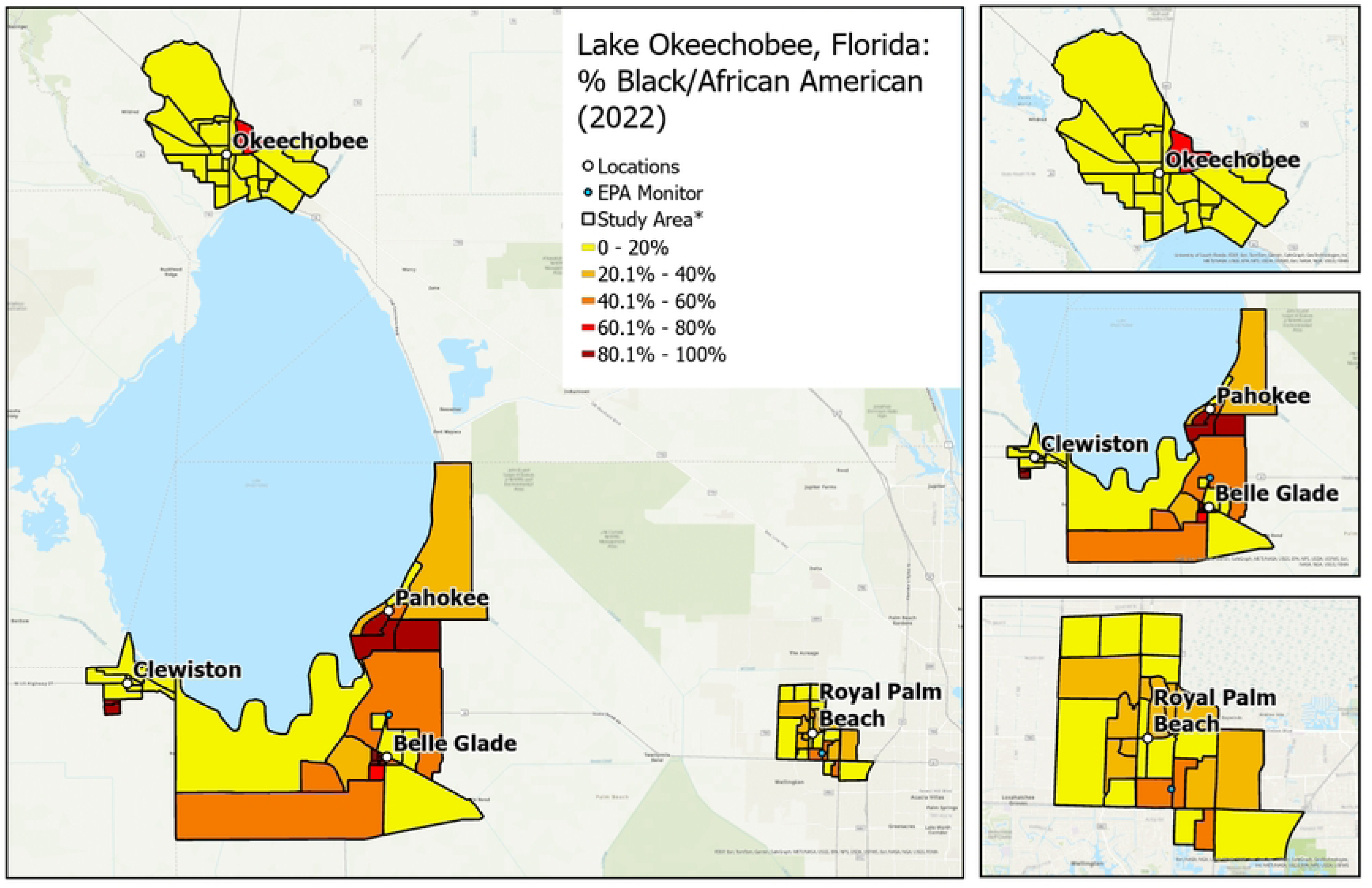
Variation in neighborhood percentage of Black/African American residents by community. * Exact locations/census block groups included will depend on recruitment.

**Figure 4.**
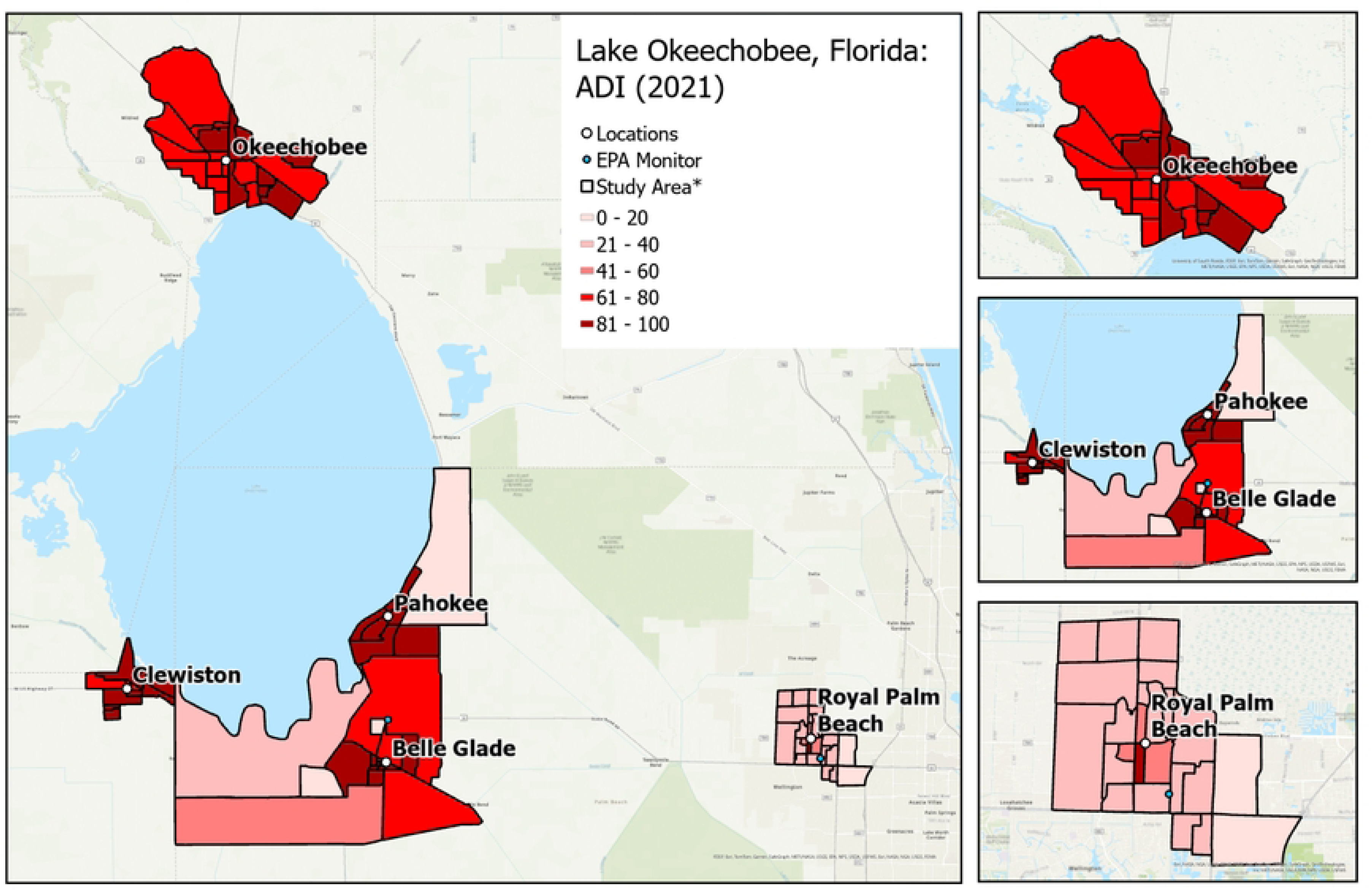
Variation in Area deprivation index (neighborhood socioeconomic status) by community. * Exact locations/census block groups included will depend on recruitment.

#### Screening and inclusion and exclusion criteria

Eligible participants will be:

- Community-dwelling individuals from the Lake Okeechobee region in Florida,
- Age 45 years and older,
- English, Spanish, or Creole speakers,
- Able to participate in study activities,
- Staying in the area for at least 12 months, and,
- Not leaving for an extended period during the summer months.

In addition to these criteria, the street address and years lived at that location will be determined. Potential participants will also be asked their date of birth (from which age is auto-calculated), their primary language, and years of education. As a means of subjective cognitive functional assessment, everyone who participates in the pre-screening will also be asked “Do you feel that your memory is getting worse?” In addition, the inclusion form will collect the town and site where the data collection will occur. Potential participants will then be pre-screened for dementia and excluded if MoCA-5 minute (Mini MoCA) scores are <12 (clinically important risk of cognitive impairment) [45]. This brief task takes ≤5 minutes and assesses the domains of attention, executive function/language, orientation, and memory. If they are unable to obtain the minimum score of 12 (adjusted for years of education), they will be referred to their provider (if unavailable, a local provider who has agreed to follow up with an assessment) and provided with a small appreciation gift for their time (writing pen) and a flyer regarding resources on brain health.

#### Recruitment strategy

Within each of the communities, the study team will conduct recruitment events at partnering churches and community and senior centers, with the goal of recruiting individuals representing the ethnoracial diversity of the chosen communities. Participants will be recruited by community research assistants (CRA) who have personal knowledge of these communities, including their places of worship, neighborhood community centers, and parent-teacher associations (several current CRAs are local educators, in addition to social workers, pastoral leaders, and other trusted gatekeepers).

Adjustments to the recruitment strategy will be made carefully and in response to opportunities that would be consistent with the study aims or due to unexpected recruitment issues. The study will be advertised via flyers and word of mouth. Enrollment will be guided by ADRD Research Recruitment principles: (1) fostering existing community partnerships, (2) building on trust to share ownership of the research mission, and (3) welcoming new partnerships based on community needs. The study embraces the principle of increasing workforce diversity and community bridges between university and community settings by hiring residents as CRAs. During participant recruitment activities, we will offer clinical measurements [e.g. HbA1C (6-month blood glucose) and body mass index (BMI)] in partnership with Healthier Glades and the local Diabetes Coalition.

#### Informed consent

If a potential participant is eligible and willing, they continue with the consent procedures using the e-consent form in REDCap. All participants receive a written copy of the consent form in their preferred language (English, Spanish, or Haitian Creole). The consent process consists of a conversation in which a bilingual member of the research team reviews the consent form, answers questions, and confirms their willingness to participate by reading the consent statement aloud. Additionally, they will be asked if they would be willing to participate in future studies. The CRA will confirm the participant’s response and indicate in REDCap that informed consent was obtained. If the enrollee affirms additional interest in participating in either the smartwatch or air pollution monitor subsample, this will also be indicated in REDCap for potential future follow-up. When subsample participants are consented, they will be asked to consent using a separate consent process with a subsample consent form which is also documented in REDCap.

#### Participant incentives

All participants will receive one $25 gift card for completing the initial research activities (enrollment and baseline data collection). Those who return for annual and follow-up visits will receive a $25 gift card after each visit. A purposive sample of 120 persons distributed throughout the five communities will be offered a $25 gift card for each month of smartwatch monitoring (2 months total). Thirty-six participants who agree to host low-cost PM_2.5_ sensors (i.e., PurpleAir monitors) monitors will receive a one-time additional gift card worth $20.

#### Timeline of study assessments

For the 36-month study, at the baseline visit (T0), which occurs immediately after consent, individuals will provide data on sociodemographics, health history, and health behaviors. Participants will return to complete six-month follow-up visits for a total of seven visits (baseline and six follow-ups). During each follow-up visit (T1-T6), individuals will complete the cognitive and mini-physical performance assessments, biomarkers (e.g., blood pressure, BMI), and PROMIS measures targeting perceptions of social isolation, alcohol use, anxiety, depression, sleep problems, physical function, fatigue, ability to participate socially, and degree of pain interference with activities of daily living. The first participant was enrolled on January 20, 2024.

For the smartwatch sub-study, participants will wear an Apple Watch SE for 2 four-week periods, one during burn season and one during a non-burn period. Social isolation and cognitive function will be assessed during both the burn and non-burn seasons to identify potential changes in response to smoke-related air pollution (PM_2.5_). These outcomes are critical for understanding the health impacts of air pollution on cognitive and social well-being. Built and social environments (e.g., park spaces and ethnoracial composition) will be characterized at baseline and repeated if more current addresses or data become available during the study. Air quality monitoring will be continuous throughout the 36-month period. Table 1 shows the schedule of assessments and instrument collection by visit.

**Table 1.**
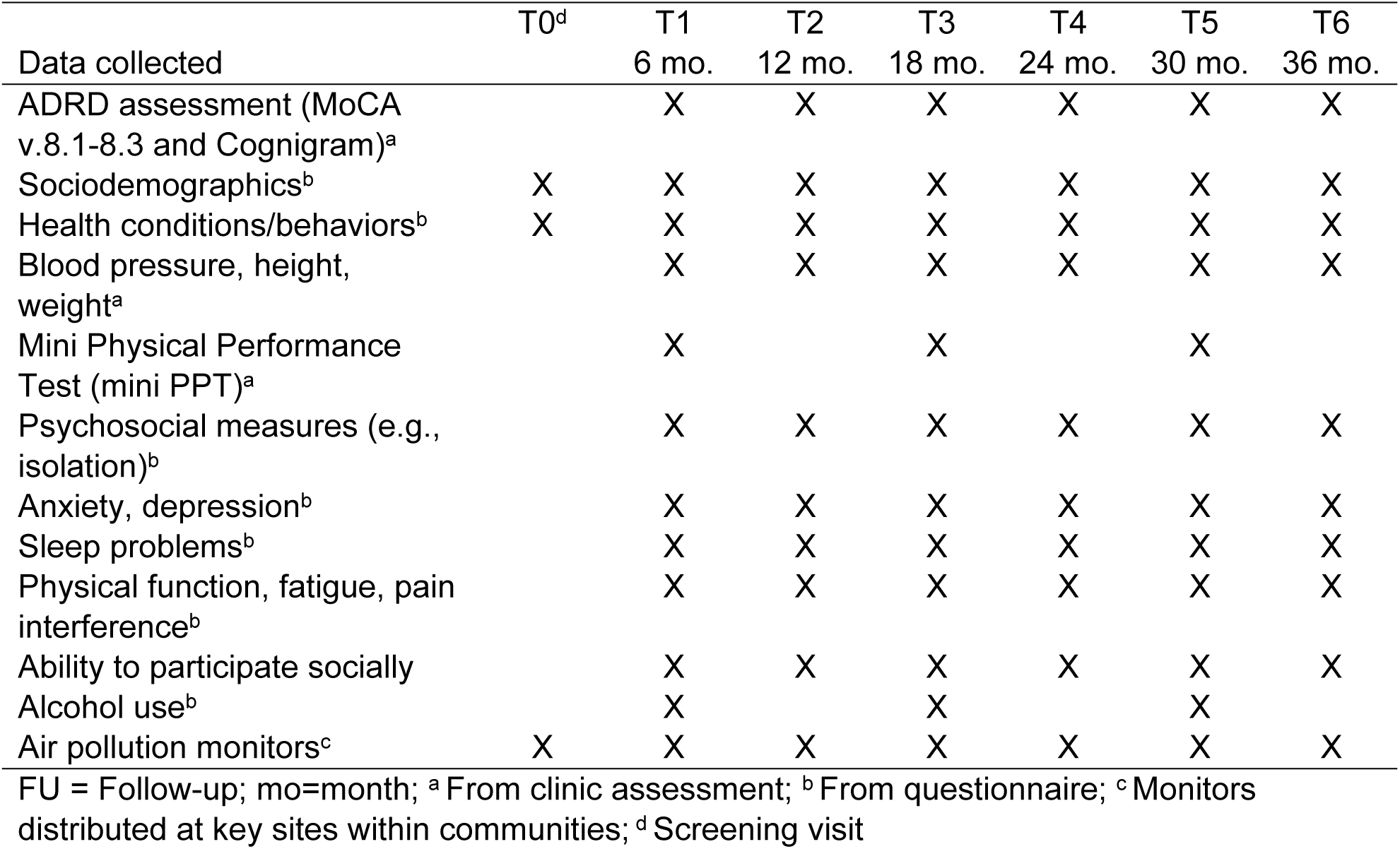
Data collection schedule for 36-month study.

### Study measures at baseline visit (T0)

#### Sociodemographics and health survey

Participants will be asked for their contact information, years living in their community, and basic demographics (e.g., gender, race/ethnicity, educational attainment, country of birth, preferred language, marital status, and religion). The survey will also ask about their household and housing/rental characteristics (e.g., living situation, number in household, home ownership/renting status, housing type, receipt of rental assistance or social security disability), their economic status (e.g., household income, employment/retirement information including whether in sugar cane fields), and primary transportation mode. Participants will be asked about their digital literacy and technology use and access (WiFi, computer, smartwatch/wearable fitness tracker). They will be asked about their health and insurance use/access and pre-existing health behaviors and conditions (i.e., smoking, asthma, chronic obstructive pulmonary disease, chronic cough, high blood pressure, heart disease, diabetes, stroke, sleep problems, head injury, hearing or vision loss, physical limitations). Lastly, participants will be asked about their physical activity frequency including how much is completed outdoors, and whether they feel safe walking in the dark in their neighborhood.

### Study measures at follow-up visits (T1-T6)

#### Sociodemographics and health survey

At the T1-T6 visits, the shortened survey will re-ask about contact information, marital status, living situation, household characteristics, employment/retirement status and details, digital literacy, technology access and use, main transportation mode and car ownership, health care and insurance use/access, smoking status, physical activity frequency, physical limitations, and history of chronic conditions. Participants will also be asked about any change in educational attainment.

#### Cognitive measures

At the T1 to T6 visits, participants will report whether they experienced worsening of their memory. Cognitive testing will include the Montreal Cognitive Assessment (MoCA) [46], a global cognition screening instrument with scores ranging from 0 to 30 with lower scores indicating greater impairment. A score of 26 or above is generally considered normal, while lower scores may indicate cognitive impairment. In addition, the Cognigram will assess four cognitive domains [psychomotor function, attention, learning, and working memory) [47, 48]. A score of < 90 on learning/working memory composite and ≥ 90 on attention/psychomotor composite indicates risk of mild cognitive impairment (MCI).

#### Physical exam measures

At the T1 to T6 visits, systolic and diastolic blood pressure and height and weight will be measured. Blood pressure will be measured by oscillometer, and high systolic blood pressure (SBP) will be defined as values ≥140 mmHg. Measured height and weight will be used to calculate BMI (kg/m^2^), with BMI values of >30kg/m^2^ indicative of obesity.

#### Modified CAIDE (mCAIDE)

At the T1, T3, and T5 visits, the mCAIDE will be calculated based on age (<65, 65-72, and >72 years), education (<12, 12-16, and >16 years), sex assigned at birth (male, female), high SBP (≥140 mmHg), obesity (>0kg/m^2^), hypercholesterolemia (yes, no), and low physical function measured via the Mini Physical Performance Test (mini PPT) (high, low). Participants will be asked if they have been told they have high total cholesterol indicating hypercholesterolemia (i.e., “Has your doctor or health professional ever told you that you have high cholesterol?”). Participants will complete the mini PPT, which requires four basic activities of daily living (i.e., picking up item from floor, 50-foot walk, sit to stand (chair rise), and progressive Rhomberg (balance) tasks). Mini PPT values ≥12 (out of 16) indicates functional physical status. The mCAIDE is a validated measure that ranges from 0 to 14 with higher scores indicative of higher risk of cognitive impairment.

#### Alcohol consumption

At the T1, T3, and T5 visits, participants will be asked about their alcohol use using the AUDIT-C (alcohol consumption), a brief, reliable, and valid screening tool for alcohol use consisting of three questions [49]. The first question “How often do you have a drink containing alcohol?” is answered on a Likert Scale from never to ≥4 times a week. If participants answer “never”, the screening ends and they receive a total score of 0. Otherwise, two additional questions will be asked about the amount of alcohol consumed: “How many on a typical day when drinking” (1 to 10 or more), and “How often do you have five or more drinks on one occasion?” answered “Never” to “Daily or almost Daily”. A score of 3 or more or reporting 6 or more drinks in a single occasion indicates risk for alcohol use disorder.

#### Psychosocial measures

At the T1 to T6 visits, the PROMIS Social Isolation 4a [50], Lubben Social Network Scale-6 (LSNS-6) [51], Berkman-Syme Social Network Index (BSNI) [52], and De Jong Gierveld Loneliness Scale will assess social isolation, social connectedness, and social networking, and loneliness, respectively [53].

- PROMIS Social Isolation 4a: A brief tool with evidence of validity and reliability (Coefficient alpha > 0.90) assessing social isolation over the past seven days with four items (e.g., feel left out) rated from 1 (Never) to 5 (Always). The total score ranges from 4 to 20, with higher scores indicating greater social isolation.
- LSNS-6: An instrument with evidence of validity and reliability (Coefficient alpha > 0.80) assessing social engagement through six items (e.g., number of relatives see/hear from at least once a month) scored from 0 to 5. The total score ranges from 0 to 30, with lower scores indicating greater social engagement.
- BSNI: An instrument with evidence of validity and reliability (Coefficient alpha = 0.79), assessing social connectedness through 11 items (e.g., frequency attending religious meetings/services) measuring four types of social connections, including marital status (married versus not), sociability (number and frequency of contacts with children, close relatives, and close friends), church group membership (yes versus no), and membership in other community organizations (yes versus no). Scores range from 0 to 4, with higher scores indicating greater connectedness.
- De Jong Gierveld Loneliness Scale: An instrument with evidence of validity and reliability (Coefficient alpha ≥ 0.70-0.76) instrument that collects 6 items (e.g., miss having people around) that measure overall loneliness, including emotional and social loneliness, with responses ranging from "*yes*" to "*more or less*" to "*no*." The total score ranges from 0 to 6, with higher scores indicating greater loneliness.

#### Anxiety and depression

At the T1-T6 visits, the PROMIS Depression-Short Form 4a [54] (Coefficient alpha > 0.90) will be used to assess depressive symptoms, with four questions (e.g., sadness and hopelessness over the past week) rated on a 5-point Likert scale ranging from 1 (Never) to 5 (Always). The total score ranges from 4 to 20, with higher scores indicating greater depressive symptoms. Also, at the T1-T6 visits, the valid and reliable (Cronbach’s a> 0.90) PROMIS Anxiety-Short Form 4a [54] will be used to assess anxiety symptoms over the past seven days. Four items (i.e., feeling uneasy) are rated from 1 (Never) to 5 (Always), with the total score ranging from 4 to 20.

#### Sleep related problems

Two instruments will be administered at visits T1 to T6 to capture sleep related problems. The PROMIS SF–Sleep Disturbance 4a [55] collects 4 items (e.g., difficulty falling asleep) on a 5-point scale (very poor to very good), with a total raw score ranging from 8 to 40. The Sleep Related Impairment 4a collects 4 items on a 5-point scale (not at all to very much), with a total raw score also ranging from 8 to 40. The raw scores for both of these are rescaled into a standardized T-score with a mean of 50 and a standard deviation of 10. Higher scores indicate a greater severity of sleep disturbance.

*Physical function, fatigue, and pain interference* will be captured at visits T1 to T6. The PROMISE SF-Physical Function 4a [56] collects 4 items (e.g., ability to do chores) on a 5-point scale (without any difficulty to unable to do), with a total score ranging from 4 to 16. The PROMISE SF-Fatigue 4a [57] collects 4 items (e.g., trouble starting things because tired) on a 5-point scale (not at all to very much), with a total score ranging from 8 to 40. The PROMISE SF-Pain Interference 4a [58] collects 4 items (e.g., how much pain interfered with chores) on a 5-point scale (not at all to very much), with a total score ranging from 6 to 30.

#### Difficulty participating socially

At visits T1 to T6, the 4-item PROMISE SF-Ability to Participate Social 4a [59] will be used to assess difficulty participating in social activity. The questions (e.g., trouble doing activities with friends) are answered on a 5-point scale (never to always) with a total raw score ranging from 4 to 20.

### Air pollution data

We will measure PM_2.5_ using two U.S. Environmental Protection Agency monitors, one located in one agricultural study community just south of Lake Okeechobee and one located in the reference community east of the main study region (Figure 1). Due to the dynamic nature (i.e., stochastic) of smoke plumes and air pollution dispersion from agricultural fires, we will also install 36 low-cost PurpleAir sensors placed in the study area by the CSU team and the community research assistants. We will ensure the air sensors are installed at residences located in majority Black/Hispanic and majority non-Hispanic White neighborhoods, according to Census data. PurpleAir monitors provide estimates of PM_2.5_ by mass using the Plantower PMS5003 laser particle counter, which does not have the accuracy of the EPA AQS monitors.

To approximate the EPA monitoring data, we (1) apply the Plantower PM_2.5_ correction factors from Barkjohn *et al.* (2021) and (2) temporally bias-correct Plantower PM_2.5_ measurements using closely located PurpleAir and EPA monitors. PurpleAir monitors are also equipped with a BOSHC BME280 to measure environmental factors (i.e., temperature, pressure, relative humidity).

All PurpleAir data from study-specific monitors are available for download through the PurpleAir Data Download Tool. Though there are not extensive publicly operating PurpleAir monitors in the study area, there are several PurpleAir monitors that operate east of the study area closer to the Atlantic coast, whichin prior study campaigns (Sablan *et al.* 2024) have been used to compare to data collected as part of our study. As of 2023, PurpleAir charges for data download for publicly operating monitors using a point-based billing system. Points range in value (100,000 to 1,000,000 per USD) depending on how many are purchased. The cost of data on a point basis depends on the data fields and number of observations requested. Based on prior work, the research team will leverage recently developed high-resolution fire-detection and Aerosol Optical Depth (AOD) products from NASA’s Moderate Resolution Imaging Radiospectrometer (MODIS), NOAA’s Visible Infrared Imaging Radiometer Suite (VIIRS), and the Geostationary Operational Environmental Satellite (GOES-16) to develop local climatologies of fire and smoke. This procedure leverages NOAA’s Hazard Mapping System (HMS) satellite-based observations to separate smoke from non-smoke influence. We will create daily kriged surfaces of bias-corrected PM_2.5_ for each study community for baseline and subsequent study years. Participants will be assigned exposures based on their home location. Based on daily estimates of PM_2.5_, we can aggregate the observations at any temporal resolution, including hour, day, week, month, and burn and non-burn seasons, to form the exposure assessment strategy for Aim 1.

### Neighborhood social and built environment variables

Neighborhood social and built environments at the residential Census block group level will be captured with 4 domain indices (green environment, neighborhood socio-economic status, social environment, walk-friendly design) and a summary Healthy Neighborhood Index (HNI). Domain categories were determined *a priori* as they are major neighborhood-level constructs associated with ADRD risk factors and disease risk in prior studies (citations). These measures will be calculated for the baseline visit year and following years, accounting for changes in residential address and changes in neighborhood environments, as applicable. ArcGIS Pro will be used to calculate and map these objective geographic information systems (GIS) measures.

#### Green environment

- Percentage of tree canopy: Tree canopy data obtained from the U.S. Forestry Service will be used to calculate the percentage of the block group comprised of tree canopy (i.e., percentage of tree coverage of the ground provided by tree leaves, branches, needles, and stems).
- Mean normalized difference vegetation index (NDVI) will be calculated for each participant’s neighborhood for the burn and non-burn periods based on satellite imagery available from the U.S. Geological Survey (USGS, Landsat 8 Level 2, Collection 2, Tier 1). ^84^ The NDVI scale is -1 to +1 (more positive measures=healthier vegetation/”greener”) and is calculated as follows: NDVI=(R_NIR_-R_RED_)/(R_NIR_+R_RED_). R_NIR_ is reflectance in near-infrared (0.86-0.88mm) and R_RED_ is reflectance in red (0.64-0.67mm). Reflectance values are captured in 30×30 meter Landsat satellite images. Mean NDVI will be calculated for participants’ Census block group. This global measure of the density of healthy vegetation captures all types of greenery including purposefully-built greenspaces (e.g., park/open spaces, street plantings, and yards) and those in natural and rural environments (e.g., forests, farms, and uninhabited areas), and has been linked to multiple ADRD risk factors and outcomes. ^65,85-87^
- Percentage open space: National Land Cover Dataset (NLCD) will be acquired through the Multi-Resolution Land Characteristics Consortium, to calculate the percentage of open space in the block group. The NLCD uses 30-meter spatial resolution satellite imagery to characterize land and surface uses and types, such as developed land, barren land, deciduous forest, scrub, and wetlands. Developed open space includes mostly vegetation/grasses (<20% impervious surfaces) found in parks, large family lots, planted vegetation for recreation or erosion control, and golf courses.
- Percentage park space, distance to nearest park, and number of parks: We will calculate the percentage of the block group comprised of park space using combined data from the Florida Geographic Data Library’s Areas of Interest layer for Florida Parks and Recreational Facilities and ESRI’s ArcGIS Online data layer “USA Parks” (covers state parks). Distance in miles from the residence to the nearest park and number of parks in the block group will be calculated based on the same park data sources.

#### Neighborhood socioeconomic status

- Area deprivation index (ADI): ADI values will be downloaded from the University of Wisconsin Neighborhood Atlas. ADI was calculated from 17 U.S. Census block group variables on socioeconomic status (e.g., median home values, % families below poverty level), representing income, education, employment, and housing quality. Higher scores (range: 1-100) equate to more socioeconomically deprived neighborhoods.
- Percentage of vacant homes, households without internet, and average monthly housing costs: Data will be obtained from the U.S. Census American Community Survey (ACS) to calculate the percentage of all housing units in the block group that are vacant and without internet, as well as the average monthly housing costs (calculated using the average of both median mortgage and median gross rent) per block group.
- Percentage of renter-occupied/for-rent homes and housing and transportation costs as a percentage of household income: Data will be obtained from the Center for Neighborhood Technology Housing and Transportation Index to calculate the percentage of all housing units that a rented/for-rent as well as the average housing plus transportation costs (as percentage of household income) in the block group.

#### Neighborhood social environment

- Crime rate: Rates of violent crimes by city and county will be downloaded from the Federal Bureau of Investigation. Violent crime includes homicide, rape, robbery, aggravated assault, property crimes, arson, burglary, larceny-theft, and motor vehicle theft.
- Neighborhood sociodemographics: U.S. Census American Community Survey data will be used to characterize participants’ block groups by their racial and ethnic composition (e.g., % Black/African American, % White, % Hispanic), percentage of residents who are >65-year-olds, the average household size, and the percentage of residents who are limited English speakers.
- The total number of churches/worship places and civil society/membership/social clubs will be calculated for each block group from parcel-level data available from the Florida Geographic Data Library (FGDL).

#### Neighborhood walk-friendly environment

- Walking destinations and percentage retail: The total number of walking destinations will be calculated for each block group using parcel data from the FGDL. These destinations will include any business or public space that could facilitate social interactions and that may promote neighborhood walking, for example a post office, bank, grocery store or restaurant. Percentage retail space for each block group will be calculated using data on retail parcels (e.g., supermarkets, auto sales, shopping malls, professional services, restaurants/cafeterias, department stores).
- Number of pedestrian or bicycle crashes per year will be determined at the city-level using University of Florida’s SIGNAL 4 Analytics program, which receives the data from the Florida Department of Highway Safety and Motor Vehicles (FLHSMV).
- Percentage sidewalk and crosswalk coverage: We will use the USGS National Transportation Dataset for road network data alongside high-resolution imagery taken from Google Earth Pro to manually delineate sidewalks next to roadways and crosswalks at intersections. For each block group, we will calculate the percentage of the roadways with sidewalks and the percentage of all intersections that include a pedestrian crosswalk.
- Maximum road speed and annual average daily traffic (AADT) data were obtained from the Florida Department of Transportation (FDOT). Mean values for maximum road speed and AADT will be calculated for the roadways within each block group.
- Intersection density: Intersection density measures (auto-oriented, 3-legged/4-legged pedestrian-oriented), available from the Environmental Protection Agency’s Smart Location Database, are calculated as the number of intersections per square mile within each Census block group. A higher intersection density correlates with a more walkable environment.
- Compact neighborhood score: The Compact Neighborhood Score will be determined for each block group from the Center for Neighborhood Technology Housing and Transportation Index data [60]. This measure highlights areas that are higher in density, mixed use, and pedestrian friendly situated closer to rail and transit stations.
- Transit Access Score and Transit Connectivity Index will be determined for each block group from the Center for Neighborhood Technology Housing and Transportation Index data, which covers over 200 metrics that help analyze the social and economic impact of transit (e.g., land area accessible from a block group within a 30 minute transit trip, the number of bus routes and train stations within walking distance for households in a block group, frequency of service for public transport).

#### Neighborhood Indices

The composite Healthy Neighborhood Index (HNI) is calculated as the average of the weighted standardized scores for the four domains or sub-indices (i.e., green environment, neighborhood SES, social environment, and walk friendly) converted to a standard normal percentile. The HNI is intended to capture the multidimensional nature of neighborhood-level factors associated with ADRD risk that cannot be explained by a single variable or a group of variables within a single domain. Data normalization is essential in composite indices as the underlying variables are often measured at different scales or units of analysis. A standard z-score calculation is used to normalize the data for the HNI composite score. Since z-score standardization requires data to be approximately normally distributed, univariate analysis is performed to identify variables with a skewed distribution and the presence of outliers. Scatter plots, box plots, or the Grubbs statistical test are employed to identify outliers. Log-10, natural log, square root, or the inverse normal method are common transformations used to create a new variable as a function f(X) of the original variable. The underlying variables for each domain or sub-index are examined for directional effect, and the z-score computation is adjusted to account for those effects. Weighting the underlying variables of each sub-index is an important consideration when different variables may have dissimilar effects on the neighborhood-level ADRD risks. In this study, we derive weights from Principal Component Analysis (PCA), specifically the component loadings matrix (post-rotation) representing the total unit ratio of explained variance for each variable loaded on a component. We compute a sub-index score for each of the four domains as the average of the underlying variables z-scores. The overall HNI index score is calculated by averaging the standard normal percentile of the four sub-components. Ordinary least squares regression analysis will be performed to gauge the internal validity of the HNI, where the HNI score will be used as the dependent variable and the four sub-indices as independent variables.

### Smartwatch sub-study data collection

Smartwatches can passively and continuously monitor a person’s routine behavior, including activity level and social interaction in real-time, using EMA responses and objective measurements, e.g. latitude, longitude. Their use in this study will provide a mechanism to model human behavior in naturalistic settings and to map digital behavior markers onto predicted clinical measures. Studying real-time data provides more detailed insights into immediate changes to behaviors and mood that occur when encountering low air quality or distressed environments and how this translates to cognitive performance.

Collected smartwatch sensor data will include accelerometer, gyroscope, heart rate, oxygen saturation, respiration rate, and location. The custom smartwatch app queries participants 4 times a day at random times during waking hours (between 8am and 8pm with a minimum one-hour gap between queries). If a user does not respond within 2 minutes, the app will repeat the query after a five-minute pause. If the user still does not respond, the app will wait until the next query time to interact with the user. The watch provides a notification in the late evening to charge the watch and in the early morning (if still on the charger) to put on the watch. We will synchronize smartwatch behavior markers and sensed location data with air quality readings and built and social environment data. Research Assistants will provide weekly check-in support for smartwatch participants.

The smartwatch measures are listed in Table 2. The daily queries ask participants to answer ecological momentary assessment (EMA) questions about their current state. Smartwatch features are extracted through a fusion of EMA, self-reports, observational health assessments, participant demographics, and selected digital behavior markers. During each query, participants will be asked Likert scale (1=Not at all to 5=Very much) questions on their physical activity, mental health status, air quality, and whether smoke in the air bothered them. In addition, during the last query of the day, participants will be asked how much time they spent with neighbors, friends, or family, and how much time they spent with others in their neighborhood, and they complete four tasks. Task 1 is the validated n-back task, which displays three shapes and asks participants to answer (yes/no) as quickly and accurately as possible whether the displayed shape is the same as the one shown before. We custom-designed a smartwatch version of the n-back task that has demonstrated reliability and validity in everyday environments [61]. The n-back task is well suited for frequent use because it is short (45 seconds) and sensitive to within-subject variations without obfuscations caused by practice effects after baseline is reached. Task 2 is an audio journal, in which participants will be asked to use the watch microphone to describe what they did that day and how they felt about their day.

**Table 2.** Schedule of assessments for smartwatch sub-study.

| Data collected | Agricultural non-burn period | Agricultural burn period |
| --- | --- | --- |
| Smartwatch sensor data: oxygen saturation, respiration rate, and location | Continuously measured for 1 month | Continuously measured for 1 month |
| EMA:<br>• In the past 30 minutes, how physically active have you been?<br>• In the past 30 minutes, how much have you felt anxious, depressed, or irritable?<br>• In the past 30 minutes, how clean has the air seemed to you?<br>• In the past 30 minutes, how much has smoke in the air bothered you?<br>• Today, how much did you spend time with neighbors, friends, or family? (only asked at last prompt)<br>• Today, how much time did you spend with others in your neighborhood? (only asked at last prompt) | 4 prompts a day, daily for 1 month | 4 prompts a day, daily for 1 month |
| Tasks:<br>• n-back<br>• Audio journal | Once a day (last prompt of the day), daily for 1 month | Once a day (last prompt of the day), daily for 1 month |

EMA = Ecological Momentary Assessment

We will then employ joint inference machine learning to predict each target variable from the combined set of features. We will report correlations between automatically derived and self-reported measures of activity and SI. To assist in ensuring a minimum 80% adherence rate, our software will alert RAs if a lapse of ≥ 1 day in data collection occurs. The RA will check in with participants each week, as well as times these lapses are detected, to answer questions about using the smartwatches.

The following statistics are calculated for each data source by day and overall: mean, standard deviation, min, max, power, zero crossing, skewness, kurtosis, peaks, autocorrelation. Data sources include the raw sensor values collected by the smartwatch. They also include the following digital markers:

- Activity level, total time for sleep, eat, work, exercise, relax, errands, travel, walk, run, lie down, based on automatically recognized activities
- Audio journal sound features: pitch, rate, intensity, spectrum, formants, harmonicity, MFCC, pauses and duration, speech beats/minute, FO
- Audio journal text features (based on speech-to-text conversion): unique phrases/words, characters/word, SMOG, named entities, sentiment/mood markers, LIWC, TF-IDF, word embeddings, LDA
- Location: distance/bearing from home, distance travelled, durations at frequented locations, transitions, air quality and temperature
- N-back: score statistics, score slope and y-intercept, convergence rate, convergence mean
- EMA: response values, response rate

### Data management plan and security

A Data Safety Management Plan is in effect and overseen by the Florida Atlantic University Data Safety Management Officer. To minimize risk of breach of confidentiality, all informed consents and study materials will be entered on encrypted, password-protected computers using an electronic REDCap database. This database will be accessible only to the authorized research team members and will be stored on secure servers at Florida Atlantic University. A dual authentication and University-issued password is required to access this database. Using brief, efficient, and culturally sensitive methods for measurement, we will take every precaution to ensure confidentiality.

### Safety considerations

A Data Safety Monitoring Board is not required for this observational cohort study. There are minimal risks associated with study measures and assessments. Data collection occurs during a one-on-one visit between the community research assistant (CRA) and participant in private settings. Embarrassment about performance, fatigue, or stress from discussion about psychosocial situations may occur. The experienced research team of healthcare professionals is prepared to deal with participant concerns and make referrals if needed. In the unlikely event that any adverse event does occur, such as a physical fall or breach of confidentiality, the FAU Institutional Review Board will be notified.

### Sample size/power

#### Sample Size Calculations for full sample

Sample size estimates were made to attain .8 power for Aims 1 and 2. The intraclass correlation was assumed to be .05 based on the cluster of neighborhoods within rural communities. For Aim 1a, an assumed effect size of *d* =.35 of the effect of PM_2.5_ on ADRD risk across burn seasons during the 3-year period was based on prior studies of cognitive decline and all-cause dementia in women related to PM_2.5_ [62]. Assuming 20 persons sampled per PM_2.5_ monitor area, this resulted in monitors covering 7 neighborhood block groups per community or N = 660 stratified across the 5 communities. For Aim 1b, effect size estimates for racial differences were based on findings of an approximately a twofold difference in hazard ratios of incident AD based on PM_2.5_ for non-Hispanic Black women compared to non-Hispanic White women or a difference in Cohen’s *d* = .45 [63] Assuming 20 persons sampled per PM_2.5_ monitor area, sample size estimates for power = .8 for the interaction of burn season by race, resulted in monitors covering at least 10 neighborhood block groups in each community for a total sample size of N = 1,000 stratified across the five communities. For Aim 2a, an assumed effect size of the built environment on cognitive function of *d* =.325 was based on Besser’s study finding of the relationship of proportion retail in 1/2-mile buffer to digit symbol coding as a measure of processing speed [64]. Assuming 20 persons sampled per neighborhood (census block), the sample size to detect an effect of the built environment on ADRD risk required six block groups per community for a total sample size of N = 600 stratified across five communities. Because Aim 1b provides the limiting sample size, the target sample size will be 1,000. Although there is a lack of literature to base the effect size for Aim 2b, the minimally detectable effect size with number of neighborhood block groups equal to 10 and n = 20 is *d* =.38. To account for 8% attrition [65–67], 1,087 participants will be recruited for the study. Conversions among risk measures and Cohen’s d were made in accordance with prior research and assumed a base rate = 0.107 of all-cause dementia [68]. Power analysis programs for mixed linear models, GLIMMPSE [69] CRT Shiny App [70] and PowerUp! [71] were used in these calculations.

#### Sample size calculations for subsample

For Aim 3, based on our prior observation of predictive correlations between smartwatch-derived behavior markers e.g. activity and clinical measures of CF i.e. MoCA) [72], we assume an effect size of clinical prediction from behavior markers of *d* = 0.52. Estimated effect size is adjusted to account for self-report error [73]. Momentary readings from a sample of N =120 is large enough to calculate 99% power for hypothesis tests. In prior studies, we found that sample sizes smaller than these provided sufficient power to evaluate learning performance [74].

### Planned statistical analyses by aim

#### Conceptual model

The statistical analysis plan is guided by our conceptual model linking neighborhood-level characteristics including PM_2.5_ exposure, social isolation, and cognitive function/ADRD risk in Figure 2.

**Figure 2.**
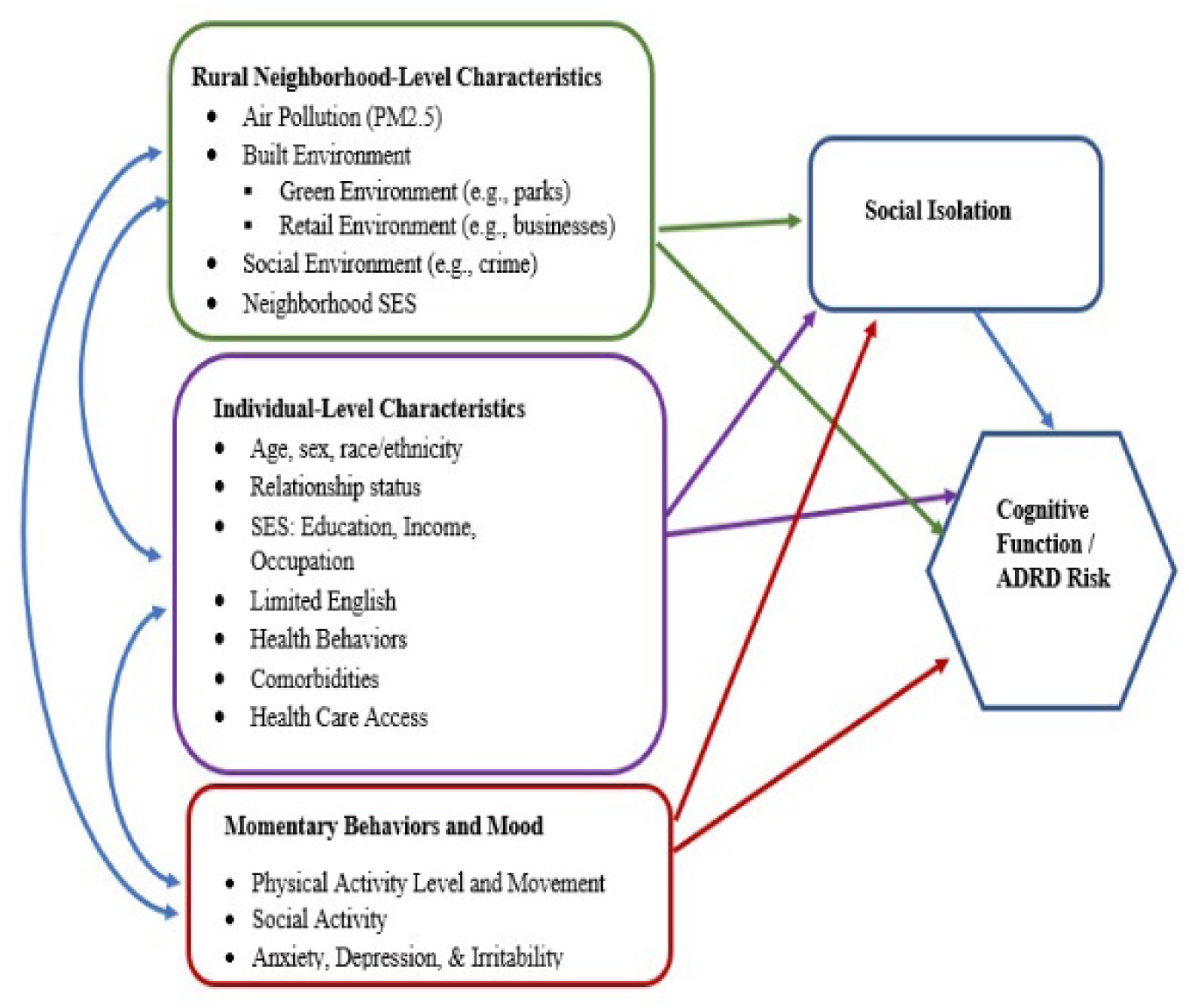
Conceptual model

#### Aim 1A

General linear mixed model (GLMM) analyses with year and season nested in participants within neighborhood block groups will be conducted for Aim 1A to evaluate the effects of changes in PM_2.5_ averaged over burn and non-burn seasons, as well as other temporal integration windows, on CF and SI. Controlling for random variance in outcomes across neighborhood block groups protects against inflated Type I errors due to neighborhood-level intraclass correlations. Additionally, linear and curvilinear trajectories for CF/ADRD risk and SI over the 3-year period associated with PM_2.5_ levels will be assessed using multilevel linear growth modeling. Both time-varying (e.g., health behaviors, work and home environments, comorbidities) and time-invariant (e.g., sex) individual- and neighborhood-level covariates (see Figure 1) will be incorporated into prediction models to control for these additional individual and neighborhood/community influences on the outcomes.

#### Aim 1B

Building on the analyses of Aim 1A, GLMM will be employed in Aim 1B to examine effect modification of Aim 1A (i.e., the differential effect of race/ethnicity on the effect of PM_2.5_ on CF and SI) through cross-level interactions of participant race/ethnicity, as well as CF and SI changes over burn season and year, controlling for neighborhood and covariate effects, as determined in Aim 1A.

#### Aim 2A

General linear mixed model analyses of participants nested within neighborhoods (i.e., census block groups) will be used to examine the effect of built and social environments on CF and SI for Aim 2A. Primary analysis will model effects of the four domains of social and built environment and the HNI on baseline CF and SI and on changes over 36 months, accounting for random neighborhood-level variance. Both time-varying (e.g., health behaviors, work and home environments, comorbidities) and time-invariant (e.g., sex) individual- and neighborhood-level covariates (see Figure 1) will be incorporated into prediction models to control for these additional individual and neighborhood/community influences on outcomes.

#### Aim 2B

Building on the analyses for Aim 2A, GLMM will be employed in Aim 2B to examine effect modification of Aim 2A (i.e., the differential effect of race/ethnicity on the effects of built and social environments on CF/ADRD risk and SI) by estimating cross-level interactions of race/ethnicity and built and social environments on the outcomes, controlling for neighborhood and covariate effects, as determined in Aim 2A.

#### Aims 3A and 3B

Random forests, convolutional neural networks, and gradient-boosted models will predict clinical measures and will be evaluated using leave-one-subject-out validation. Performance will be compared with mean and median baseline predictions for statistical significance. The primary analysis will model effects of activity level, social activity, and built environment averaged over burn season and year, although changes over time will also be assessed. Based on a continuous collection of over 8 weeks from 120 participants, we will collect at least 1.5 billion movement readings, 625,000 locations, 43,000 EMA responses, and 3,000 n-back scores. We will use the EMA responses, n-back scores, and ADRD risk scores as our target variables. The independent variables used to form the machine learning feature vectors will be daily digital behavior markers for each participant.

### Ethical considerations and declarations

The RRR study is funded by a U.S. National Institute on Aging grant (R01AG083925). The planned human subjects research has been approved via Florida Atlanta University’s (FAU) Institutional Review Board (IRB), which is the single IRB (sIRB) that is relied upon by the collaborating institutions (University of Miami, Washington State University, University of Arizona, and Colorado State University).

### Dissemination and data sharing plan

Study findings will be shared with the participants and communities under study via community presentations and will be disseminated more broadly via peer-reviewed publications and local, national, and international conference presentations. Data will be aggregated/summarized when sharing findings and no identifying information will be revealed.

We will share all exposure data (e.g., PM_2.5_ exposure data) by depositing these data with Mountain Scholar, a free-accessible repository collectively supported by institutions of higher education in Colorado and Wyoming (https://lib.colostate.edu/find/csu-digital-repository/). These data are accessible via digital object identifier and are open access; for example, the website for the preliminary exposure data for our study can be found at http://dx.doi.org/10.25675/10217/193258. This doi will be included in all of our presentations and publications so that outside research groups are made aware of this resource for use and can use this resource for independent studies.

## DISCUSSION

The RRR cohort study will follow a diverse Florida cohort from predominantly rural communities over 36 months to examine the impact of smoke-related ambient PM_2.5_ exposure and built and social environments on social isolation and cognitive decline. The Lake Okeechobee area provides a fitting setting to carry out the study aims due to its rare combination of ethnoracial diversity, greater neighborhood disadvantage, rural locale, and proximity to agricultural burning. Our study will be innovative in employing smartwatches and EMA over a two-month period, during burn and non-burn seasons, to assess momentary changes in behavioral markers, mood, and cognitive performance associated with changes in air quality and the social and built environment.

The study has several strengths, including its large sample size and recruitment from numerous block groups across South Central Florida, which ensures a range of PM_2.5_ and social and built environment exposures. Participants will be able to complete assessments in Spanish, English, or Haitian Creole, helping to ensure representation of the diverse residents living in the rural communities. We will use PM_2.5_ data sources including deployed PurpleAir monitors to support more accurate estimation of temporal smoke-related PM_2.5_ exposure. We will employ CRAs who are knowledgeable about the rural communities in the study, embracing the principle of increasing workforce diversity and community bridges between university and community settings and the “What Matters Most” caring science framework [75]. CRAs will contact participants bimonthly via phone or in-person visits to aid in retention. Despite these strengths, the study also has some limitations. Generalizability may be limited to diverse farming communities who often experience large health disparities. In addition, it is possible that we may experience trouble recruiting from some rural communities despite the Principal Investigator’s vast network of established collaborators in the study area. To combat this, we will constantly monitor recruitment numbers by community and will adjust our recruitment sources and strategies as necessary (e.g., working with faith-based leaders in those communities to determine alternative locations and organizations for recruitment and partnership).

This groundbreaking study will investigate the social and cognitive impacts of smoke-related PM_2.5_ and social and built environments in rural and ethnoracially diverse populations. While international studies have demonstrated health effects of agricultural burning, studies are limited in the U.S., and this would be one of the first U.S. studies to investigate agricultural burning in relation to cognitive health. Altogether, our findings will contribute significantly to the limited literature on this topic, identifying potential synergistic effects of individual and community characteristics on brain health inequities in a unique, rural setting of the U.S.

## Data Availability

No datasets were generated or analysed during the current study. All relevant data from this study will be made available upon study completion.

## Acknowledgements

This study is supported by the National Institutes of Health/National Institute on Aging (R01AG083925). We express our immense gratitude to our community partners and Research Assistants.

